# Genetic correlation and causality between smoking and 42 neuropsychiatric and gastrointestinal diseases

**DOI:** 10.1101/2024.05.09.24307140

**Authors:** Jiayi Xiong, Zheng Wang, Yuanfeng Huang, Shiyu Zhang, Guang Yang, Jiaqi Yang, Shuo Gao, Tianyang Wang, Jinchen Li, Guihu Zhao, Bin Li

## Abstract

**Background:** Previous investigations have elucidated epidemiological associations linking smoking to neuropsychiatric and gastrointestinal diseases, yet the underlying causal relationships remain enigmatic. To shed light on this matter, we undertook a Mendelian randomisation(MR) study with the aim of gauging the potential causal association between smoking and the susceptibility to neuropsychiatric and gastrointestinal tract diseases.

**Methods:** We meticulously collected and preprocessed genome-wide association study (GWAS) data encompassing smoking (280,508 cases and 180,558 controls) as well as neuropsychiatric and gastrointestinal phenotypes (n = 6,681 to 87,3341). To investigate the genetic correlation between smoking and diseases, we employed linkage disequilibrium score regression. We further applied multi-trait analysis of GWAS to identify the shared risk single-nucleotide polymorphisms (SNPs) implicated in both smoking and diseases. Pleiotropic genes were annotated by enrichment analysis. Subsequently, bidirectional MR analysis was performed to infer causality.

**Results:** Our findings, supported by robust evidence derived from an expansive sample size, demonstrate that smoking exerts a causal influence on merely six of these diseases, while no disease was found to causally impact smoking. Intriguingly, we discovered 513 pleiotropic genes enriched in pathways such as the regulation of growth and synapses, suggesting a potential shared genetic basis between smoking and these diseases, leading to aberrant neural development. Remarkably, among the 42 diseases scrutinized, a significant genetic correlation was exclusively observed with gastroesophageal reflux disease (GRED). Furthermore, we identified risk SNPs shared by smoking and GRED.

**Conclusions:** This study revealed the shared genetic basis and causal effects connecting smoking to neuropsychiatric and gastrointestinal diseases, thereby providing novel etiological insights into the role of smoking in these diseases.

## Background

Tobacco utilization constitutes a momentous global public health concern and stands as a paramount contributor to preventable mortality(1). Previous epidemiological inquiries have surfaced, underscoring smoking as a risk factor for many diseases, including cancers, cardiovascular diseases, chronic obstructive pulmonary disease and so on(2). In recent years, the advent of genome-wide association studies (GWAS) has engendered a burgeoning body of evidence, illuminating the shared genetic underpinnings between smoking and various diseases. Genetic correlation analysis, predicated on GWAS summary statistics, has unveiled the genetic intertwinement linking smoking to metabolic disorders, autoimmune conditions, psychiatric disorders and nervous system diseases(3–5). Beyond genetic correlation, methodologies such as Mendelian randomisation (MR), which uses genetic variants as instrumental variables, have been employed to discern the causal connections between lifestyle choices and diseases(6). Previous MR endeavors have provided insights into the causal ramifications of smoking on diseases such as lung cancer, coronary heart disease, and type 2 diabetes(7–9).

It is firmly established that smoking bears an association with neuropsychiatric and gastrointestinal diseases, both of which bear an intimate nexus with the brain-gut axis(10,11). The brain-gut axis epitomizes the bidirectional communication interlinking the central nervous system and the gastrointestinal tract, constituting a pivotal arbiter for homeostasis(10). Perturbations within the purview of the brain-gut axis have been inextricably implicated in an assortment of ailments. For instance, patients afflicted with irritable bowel syndrome have evinced perturbed interactions within the brain-gut axis(12). Although smoking has been linked to both types of diseases, including Parkinson’s disease(13) and colorectal cancer(14), in epidemiological studies, the extent of genetic correlation and causation between smoking and these diseases remains largely unknown.

This pioneering study represents an all-encompassing inquiry, scrutinizing the genetic correlation between smoking and 42 neuropsychiatric and gastrointestinal diseases. Furthermore, we unveiled the shared genetic substratum and the potential causative associations. Our approach embraces an expansive and comprehensive sample size, encompassing a wide spectrum of diseases, thereby fomenting a holistic comprehension of the mechanistic role played by smoking. This distinguishing facet sets our investigation apart from previous research, emphasising the profound genetic insights it yields.

## Methods

### 2.1 Study overview and data resources

Briefly, this study obtained GWAS summary statistics for smoking and 42 neuropsychiatric and gastrointestinal diseases. The GWAS summary statistics for smoking and the diseases were sourced from publicly available databases. Specifically, the smoking GWAS data originated from a meta-analysis encompassing the UK Biobank and the Tobacco and Genetics Consortium datasets, focusing on smoking status (ever versus never smoked). This dataset included 280,508 individuals who were ever smokers and 180,558 individuals who were never smokers, all of European ancestry. The data were obtained from the esteemed MRC-IEU OpenGWAS database, with the accession ID: ukb-b-20261(accessible at https://gwas.mrcieu.ac.uk/). The GWAS data pertaining to the 42 diseases were acquired from the MRC-IEU OpenGWAS database, the Psychiatric Genomics Consortium (PGC), and the GWAS catalog, ensuring a sample size exceeding 5,000. Detailed summary statistics for the GWAS data employed in this study can be found in Table 1.

**Table 1.**
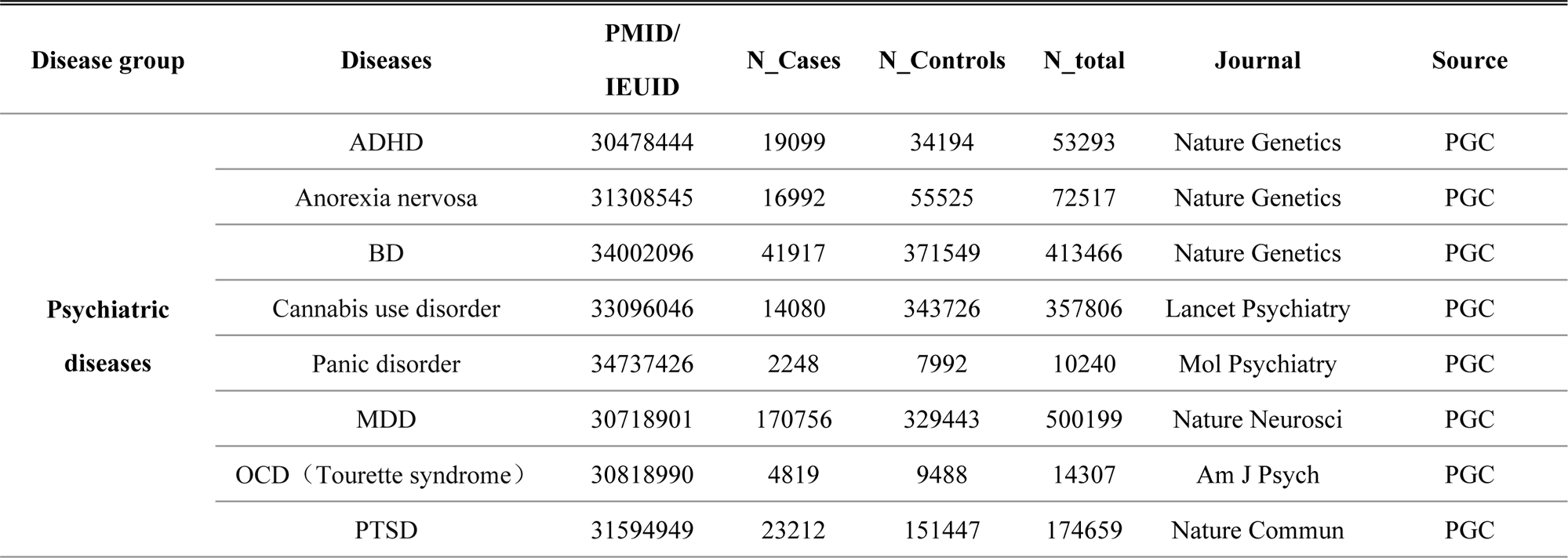
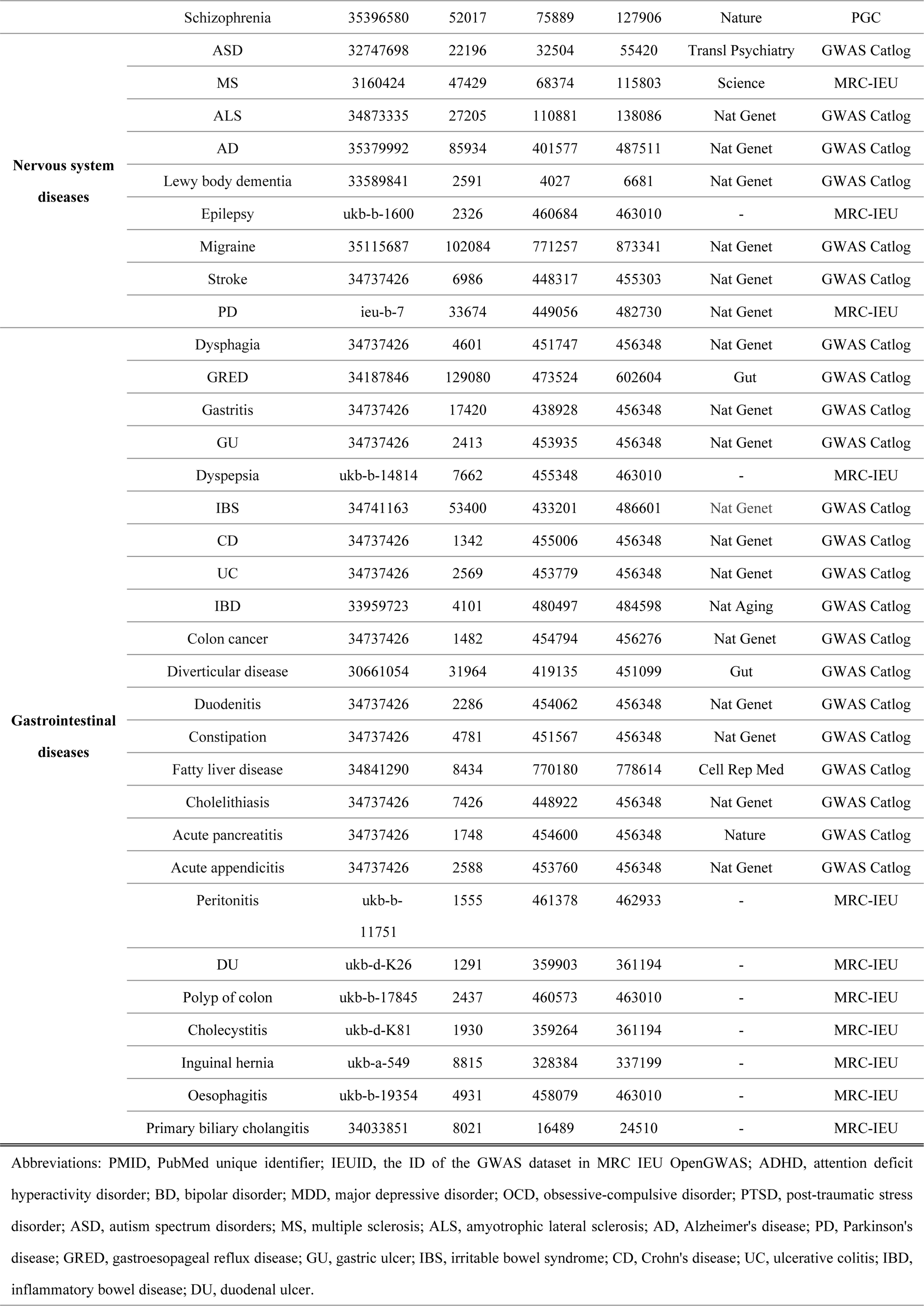
Details of GWAS Summary Data Sources.

### 2.2 Statistical Analysis

#### 2.2.1 Genetic correlation analysis

To explore the genetic correlation between smoking and the aforementioned 42 diseases, we employed linkage disequilibrium score regression (LDSC)(15). This method involves regressing the product of z scores from two GWAS scans against a measure of linkage disequilibrium for each single nucleotide polymorphism (SNP). The genetic correlation results were subjected to correction for multiple testing using the rigorous Bonferroni correction, ensuring the statistical integrity of these findings. Genetic correlations with a P-value below the significance threshold (0.05/42 = 1.25×10^−3^) were deemed statistically significant.

#### 2.2.2 Pleiotropy analysis

For diseases exhibiting significant genetic correlation with smoking according to LDSC analysis, we conducted multitrait analysis of GWAS (MTAG)(16,17) to identify shared pleiotropic SNPs between smoking and the respective diseases. MTAG is an advanced statistical fine-mapping method that leverages summary statistics from multiple GWAS to enhance the power of detecting SNPs associated with traits. It models the effect sizes of each SNP on different traits, assuming a distribution with a variance‒covariance matrix proportional to the genetic correlation matrix. MTAG was executed with default parameters, and the genome-wide significance threshold was set at P<5×10^−8^. The results include a residual correlation matrix between estimated traits, a genetic correlation matrix, and association statistics for each SNP.

To discern pleiotropic genes, we employed MAGMA(18), a gene and pathway analysis tool grounded in a multiple linear principal components regression model utilizing GWAS summary statistics. The reference panel employed was the linkage disequilibrium structure derived from European ancestry samples of the 1000 Genomes Project Phase 3. Genes exhibiting a false discovery rate (FDR)-corrected P value below 0.05 in both smoking and at least one disease were defined as pleiotropic genes.

Pathway enrichment analysis of the identified pleiotropic genes was performed using Metascape(19). The enriched Gene Ontology (GO) terms were visually represented using the clusterProfiler(20) R package.

#### 2.2.3 Causal inference analysis

To infer potential causal relationships between smoking and the diseases under investigation, we conducted bidirectional MR analysis using the R package TwoSampleMR(21). The GWAS summary statistics for smoking served as the exposure variable, while each disease was considered an outcome variable. Five different MR methods, including inverse variance weighted (IVW), MR‒Egger, weighted median, simple mode, and weighted mode methods, were adopted. The IVW method was selected as the primary approach due to its high efficiency and minimal assumptions(22). SNPs with an F statistic greater than 10 were included as instrumental variables. The MR results were adjusted for multiple testing using the FDR method. Causal associations were reported solely for analysis without significant heterogeneity or horizontal pleiotropy, as determined by both Cochran’s Q test and the MR-PRESSO test. For a significant causal relationship, we conducted a leave-one-out analysis to assess whether the results were driven by any individual SNP.

## Results

### 3.1 Summary information for GWAS data

The GWAS summary statistics used in this study are presented in Table 1. The smoking GWAS data encompassed 280,508 individuals classified as ever smokers and 180,558 individuals classified as never smokers of European ancestry, acquired from the MRC-IEU OpenGWAS database (ID: ukb-b-20261). The GWAS data for the 42 neuropsychiatric and gastrointestinal diseases were derived from individuals of European ancestry with sample sizes greater than 5,000 (n = 6,681 to 87,3341). (Table 1)

### 3.2 Genetic correlation analysis

The genetic correlation results between smoking and these 42 diseases estimated by LDSC are shown in Table 2. Following Bonferroni correction, only one disease, gastroesophageal reflux disease (GRED), exhibited a statistically significant negative genetic correlation with smoking (rg = −0.226, P = 1.27×10^−7^). The negative genetic correlation suggests the existence of shared genetic factors between smoking and GRED, albeit with an inverse association. This implies that certain genetic variants associated with smoking may confer a protective effect against the development of GRED. This discovery holds valuable implications for our understanding of the genetic determinants underlying smoking behavior and its potential impact on disease susceptibility. Further investigation is warranted to unravel the precise mechanisms underlying this relationship and explore potential therapeutic avenues for GRED.

**Table 2.**
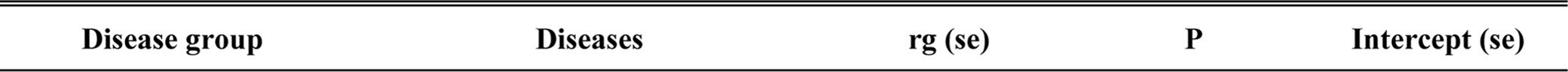
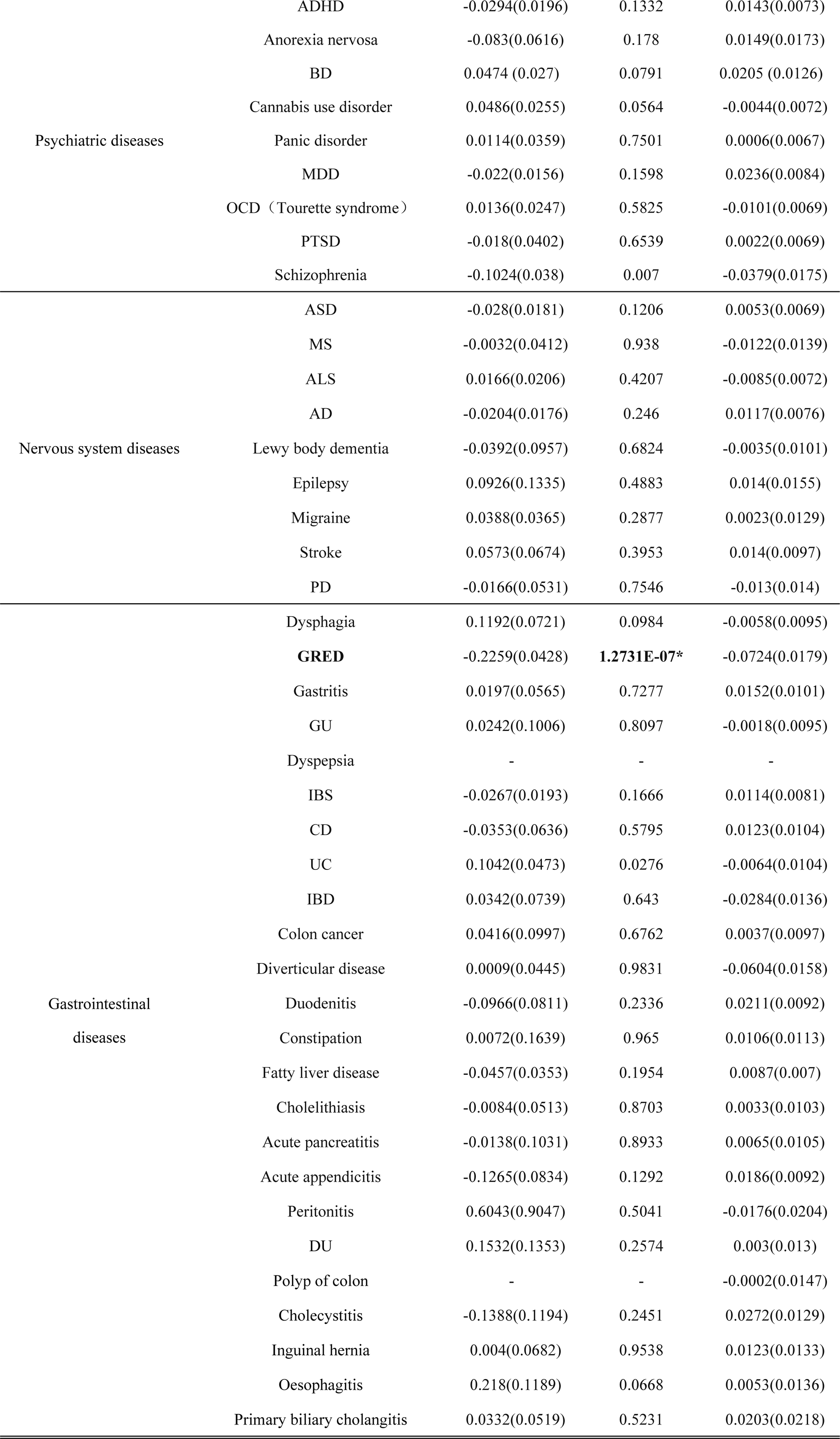
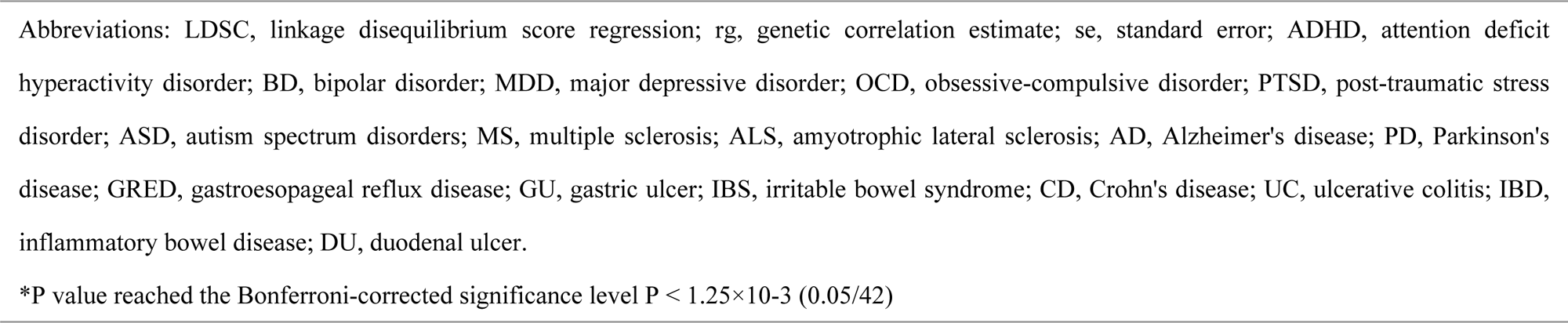
Genetic Correlation Between “ever smoked” and 42 Diseases Estimated by LDSC.

| Disease group | Diseases | $r_g$ (se) | P | Intercept (se) |
| --- | --- | --- | --- | --- |
| Psychiatric diseases | ADHD | -0.0294(0.0196) | 0.1332 | 0.0143(0.0073) |
|  | Anorexia nervosa | -0.083(0.0616) | 0.178 | 0.0149(0.0173) |
|  | BD | 0.0474 (0.027) | 0.0791 | 0.0205 (0.0126) |
|  | Cannabis use disorder | 0.0486(0.0255) | 0.0564 | -0.0044(0.0072) |
|  | Panic disorder | 0.0114(0.0359) | 0.7501 | 0.0006(0.0067) |
|  | MDD | -0.022(0.0156) | 0.1598 | 0.0236(0.0084) |
|  | OCD (Tourette syndrome) | 0.0136(0.0247) | 0.5825 | -0.0101(0.0069) |
|  | PTSD | -0.018(0.0402) | 0.6539 | 0.0022(0.0069) |
|  | Schizophrenia | -0.1024(0.038) | 0.007 | -0.0379(0.0175) |
| Nervous system diseases | ASD | -0.028(0.0181) | 0.1206 | 0.0053(0.0069) |
|  | MS | -0.0032(0.0412) | 0.938 | -0.0122(0.0139) |
|  | ALS | 0.0166(0.0206) | 0.4207 | -0.0085(0.0072) |
|  | AD | -0.0204(0.0176) | 0.246 | 0.0117(0.0076) |
|  | Lewy body dementia | -0.0392(0.0957) | 0.6824 | -0.0035(0.0101) |
|  | Epilepsy | 0.0926(0.1335) | 0.4883 | 0.014(0.0155) |
|  | Migraine | 0.0388(0.0365) | 0.2877 | 0.0023(0.0129) |
|  | Stroke | 0.0573(0.0674) | 0.3953 | 0.014(0.0097) |
|  | PD | -0.0166(0.0531) | 0.7546 | -0.013(0.014) |
| Gastrointestinal diseases | Dysphagia | 0.1192(0.0721) | 0.0984 | -0.0058(0.0095) |
|  | **GRED** | -0.2259(0.0428) | **1.2731E-07\*** | -0.0724(0.0179) |
|  | Gastritis | 0.0197(0.0565) | 0.7277 | 0.0152(0.0101) |
|  | GU | 0.0242(0.1006) | 0.8097 | -0.0018(0.0095) |
|  | Dyspepsia | - | - | - |
|  | IBS | -0.0267(0.0193) | 0.1666 | 0.0114(0.0081) |
|  | CD | -0.0353(0.0636) | 0.5795 | 0.0123(0.0104) |
|  | UC | 0.1042(0.0473) | 0.0276 | -0.0064(0.0104) |
|  | IBD | 0.0342(0.0739) | 0.643 | -0.0284(0.0136) |
|  | Colon cancer | 0.0416(0.0997) | 0.6762 | 0.0037(0.0097) |
|  | Diverticular disease | 0.0009(0.0445) | 0.9831 | -0.0604(0.0158) |
|  | Duodenitis | -0.0966(0.0811) | 0.2336 | 0.0211(0.0092) |
|  | Constipation | 0.0072(0.1639) | 0.965 | 0.0106(0.0113) |
|  | Fatty liver disease | -0.0457(0.0353) | 0.1954 | 0.0087(0.007) |
|  | Cholelithiasis | -0.0084(0.0513) | 0.8703 | 0.0033(0.0103) |
|  | Acute pancreatitis | -0.0138(0.1031) | 0.8933 | 0.0065(0.0105) |
|  | Acute appendicitis | -0.1265(0.0834) | 0.1292 | 0.0186(0.0092) |
|  | Peritonitis | 0.6043(0.9047) | 0.5041 | -0.0176(0.0204) |
|  | DU | 0.1532(0.1353) | 0.2574 | 0.003(0.013) |
|  | Polyp of colon | - | - | -0.0002(0.0147) |
|  | Cholecystitis | -0.1388(0.1194) | 0.2451 | 0.0272(0.0129) |
|  | Inguinal hernia | 0.004(0.0682) | 0.9538 | 0.0123(0.0133) |
|  | Oesophagitis | 0.218(0.1189) | 0.0668 | 0.0053(0.0136) |
|  | Primary biliary cholangitis | 0.0332(0.0519) | 0.5231 | 0.0203(0.0218) |

Abbreviations: LDSC, linkage disequilibrium score regression; rg, genetic correlation estimate; se, standard error; ADHD, attention deficit hyperactivity disorder; BD, bipolar disorder; MDD, major depressive disorder; OCD, obsessive-compulsive disorder; PTSD, post-traumatic stress disorder; ASD, autism spectrum disorders; MS, multiple sclerosis; ALS, amyotrophic lateral sclerosis; AD, Alzheimer's disease; PD, Parkinson's disease; GRED, gastroesophageal reflux disease; GU, gastric ulcer; IBS, irritable bowel syndrome; CD, Crohn's disease; UC, ulcerative colitis; IBD, inflammatory bowel disease; DU, duodenal ulcer.
\*P value reached the Bonferroni-corrected significance level $P < 1.25 \times 10^{-3}$ (0.05/42)

### 3.2 Pleiotropy analysis

Given that only GRED showed a significant genetic correlation with smoking, we proceeded to perform MTAG between the GWAS data for smoking and GRED. A total of 971,487 SNPs were included in the meta-analysis by MTAG. Among them, 358 SNPs reached the genome-wide significance level (P < 5×10^−8^) in the MTAG smoking analysis, indicating their status as pleiotropic SNPs shared by smoking and GRED (detailed information provided in Additional file: Supplementary Table S1).

Using MAGMA, we identified 513 pleiotropic genes associated with both smoking and at least one disease after excluding genes on chromosome 6 (Additional file: Supplementary Table S2). These genes are considered candidate pleiotropic genes that are shared between smoking and multiple diseases, indicating their potential role in modulating the relationship between smoking and these diseases. Notably, genes such as CRHR1, KANSL1, PLEKHM1, STH and WNT3 on chromosome 17 as well as MSRA on chromosome 8 were identified as pleiotropic genes in at least 5 diseases respectively.

Pathway enrichment analysis of the identified pleiotropic genes revealed significant enrichment in multiple GO terms (Figure 1; Additional file: Supplementary Table S3-S5), encompassing processes such as cell projection morphogenesis, presynapse, and protein domain specific binding. Importantly, our findings indicate that smoking interacts with these signaling pathways, contributing to the occurrence of 42 neuropsychiatric and gastrointestinal diseases. Remarkably, our results align with previous literature, substantiating the consistent relationship between smoking and these diseases at a molecular level.

**Fig. 1.**
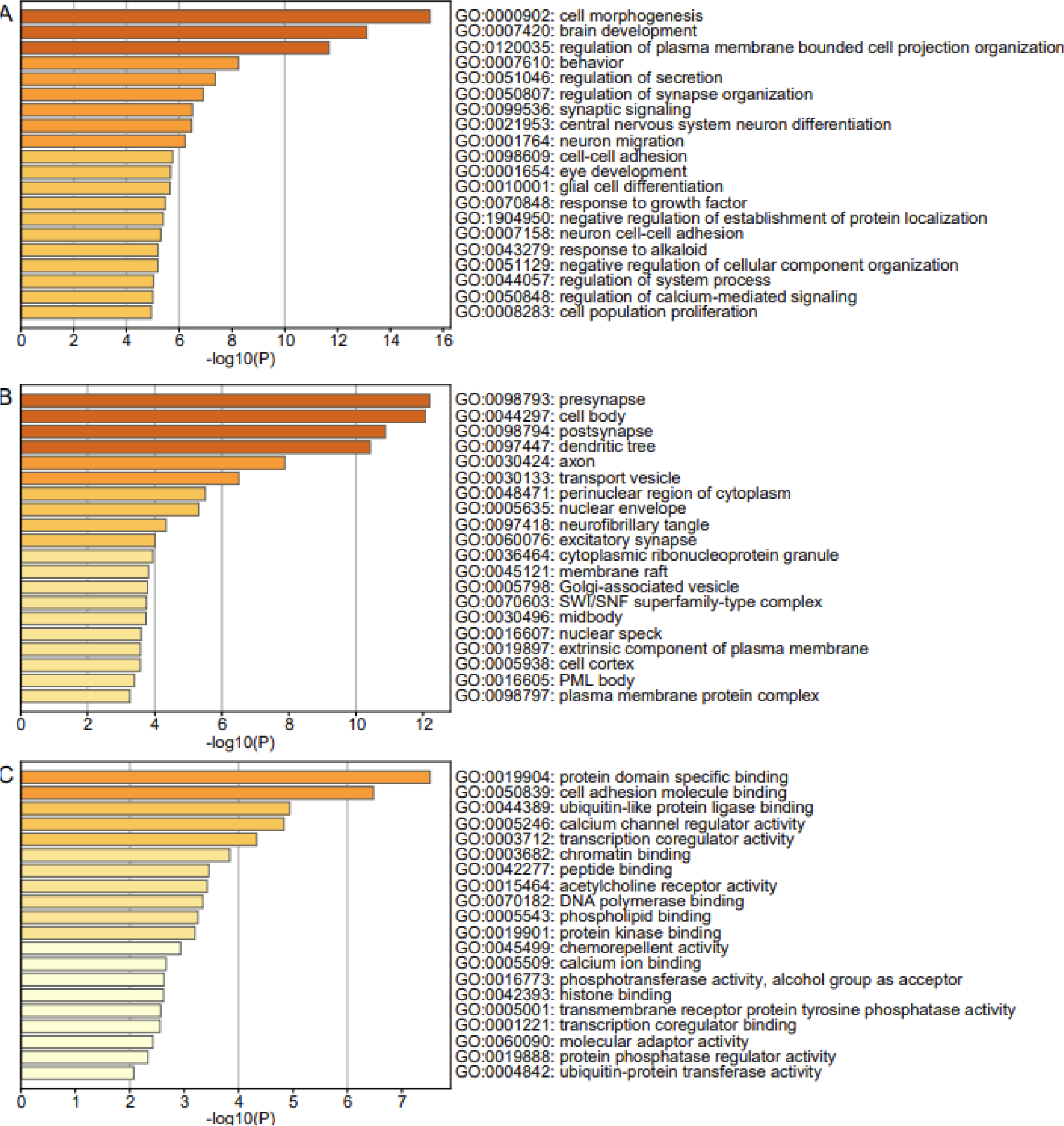
Enrichment maps of pleiotropic genes for smoking and 42 diseases. **A** GO biological process enrichment bar chart. **B** GO cellular component enrichment bar chart. **C** GO molecular function enrichment bar chart. From panels **A** to **C**, the *X*-axis is –log10(P), and the *Y*-axis is the GO term. The colour represents the enriched significance.

### 3.3 Causal inference analysis

Mendelian randomisation analysis suggested potential causal relationships between smoking and several diseases. Overall, smoking demonstrated significant positive causal effects on 6 diseases after adjusting for multiple testing using the Benjamini‒Hochberg (BH) method for P value adjustment (Figure 2; Additional file: Supplementary Table S6), including ulcerative colitis (UC) (IVW: odds ratio [OR] = 6.84, 95% confidence interval [CI] 2.23-20.99, P < 0.001), inflammatory bowel disease (IBD) (IVW: OR = 1.01, 95% CI 1.00-1.02, P = 0.003), gastritis (IVW: OR = 1.79, 95% CI 1.13-2.82, P = 0.013), post-traumatic stress disorder (PTSD) (IVW: OR = 1.97, 95% CI 1.14-3.41, P = 0.016), cholelithiasis (IVW: OR = 2.03, 95% CI 1.08-3.80, P = 0.027) and epilepsy (IVW: OR = 1.01, 95% CI 1.00-1.01, P = 0.035). Although Crohn’s disease (CD) and primary biliary cholangitis did not reach the 0.05 significance threshold, they also exhibited positive causal tendencies. It is important to note that while smoking showed positive causal effects on attention-deficit/hyperactivity disorder (ADHD), cannabis use disorder, major depressive disorder (MDD), bipolar disorder (BD), and GRED, further verification is required for this causal relationship, as indicated by the heterogeneity test and MR-PRESSO test P values being <0.05, suggesting the presence of pleiotropy and less reliable MR results.

**Fig. 2.**
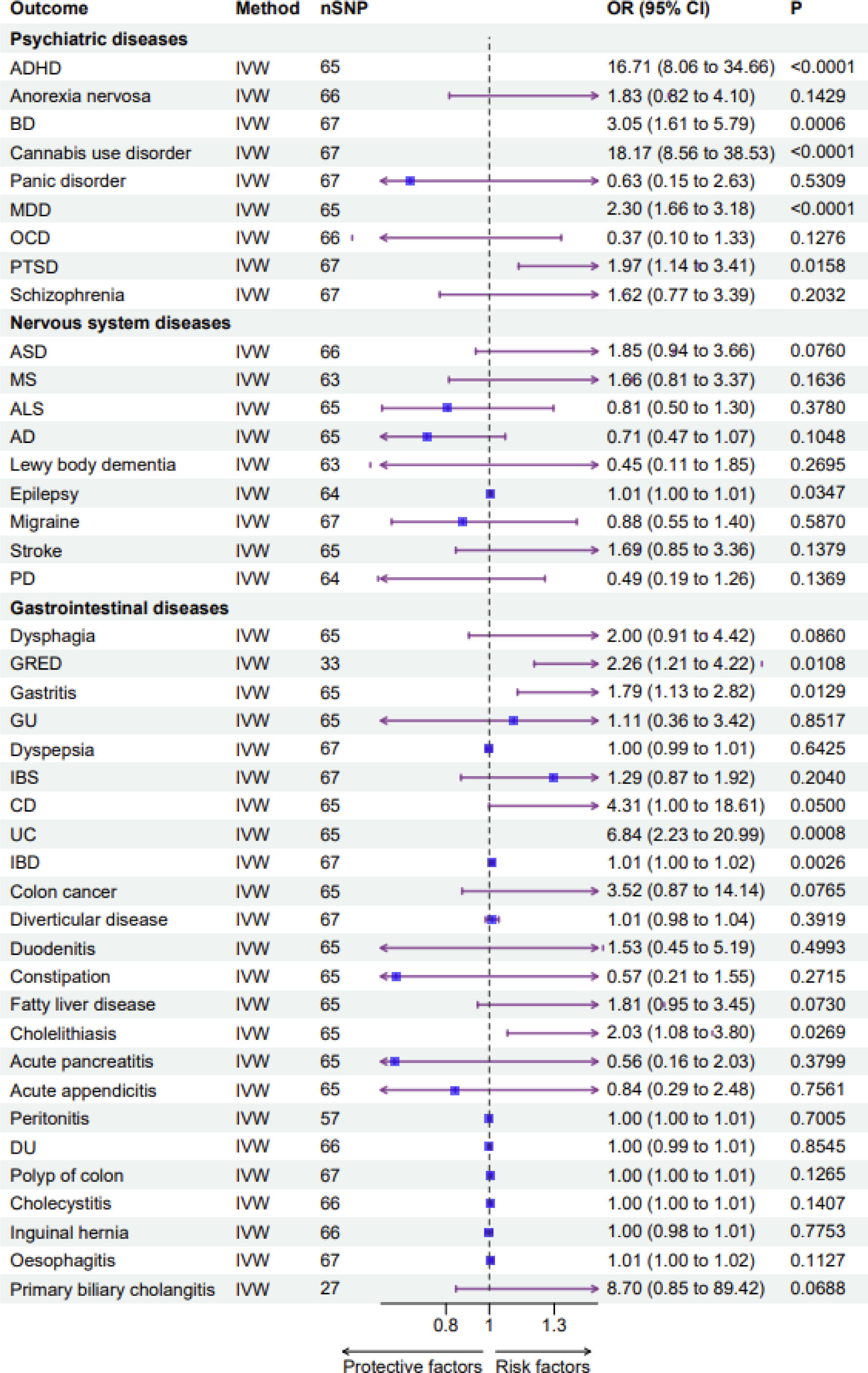
MR estimates for the effect of “ever smoked” on 42 diseases. Estimates were derived by the inverse-variance weighted method. Reported P-values were adjusted for multiple testing using the Benjamini-Hochberg procedure. The solid lines represent 95% CI; CI, confidence interval; OR, odds ratio. ADHD, attention deficit hyperactivity disorder; BD, bipolar disorder; MDD, major depressive disorder; OCD, obsessive-compulsive disorder; PTSD, post-traumatic stress disorder; ASD, autism spectrum disorders; MS, multiple sclerosis; ALS, amyotrophic lateral sclerosis; AD, Alzheimer’s disease; PD, Parkinson’s disease; GRED, gastroesopageal reflux disease; GU, gastric ulcer; IBS, irritable bowel syndrome; CD, Crohn’s disease; UC, ulcerative colitis; IBD, inflammatory bowel disease; DU, duodenal ulcer.

On the other hand, no disease demonstrated a significant causal effect on smoking after adjusting for multiple testing (Figure 3; Additional file: Supplementary Table S7). Although BP (IVW: OR = 0.99, 95% CI 0.98-1.00, P = 0.018) presented a negative causal relationship with smoking, in the MR analysis, the result was less reliable due to the detection of horizontal pleiotropy. Leave-one-out analysis indicated that the causal relationships were not driven by any single SNP.

**Fig. 3.**
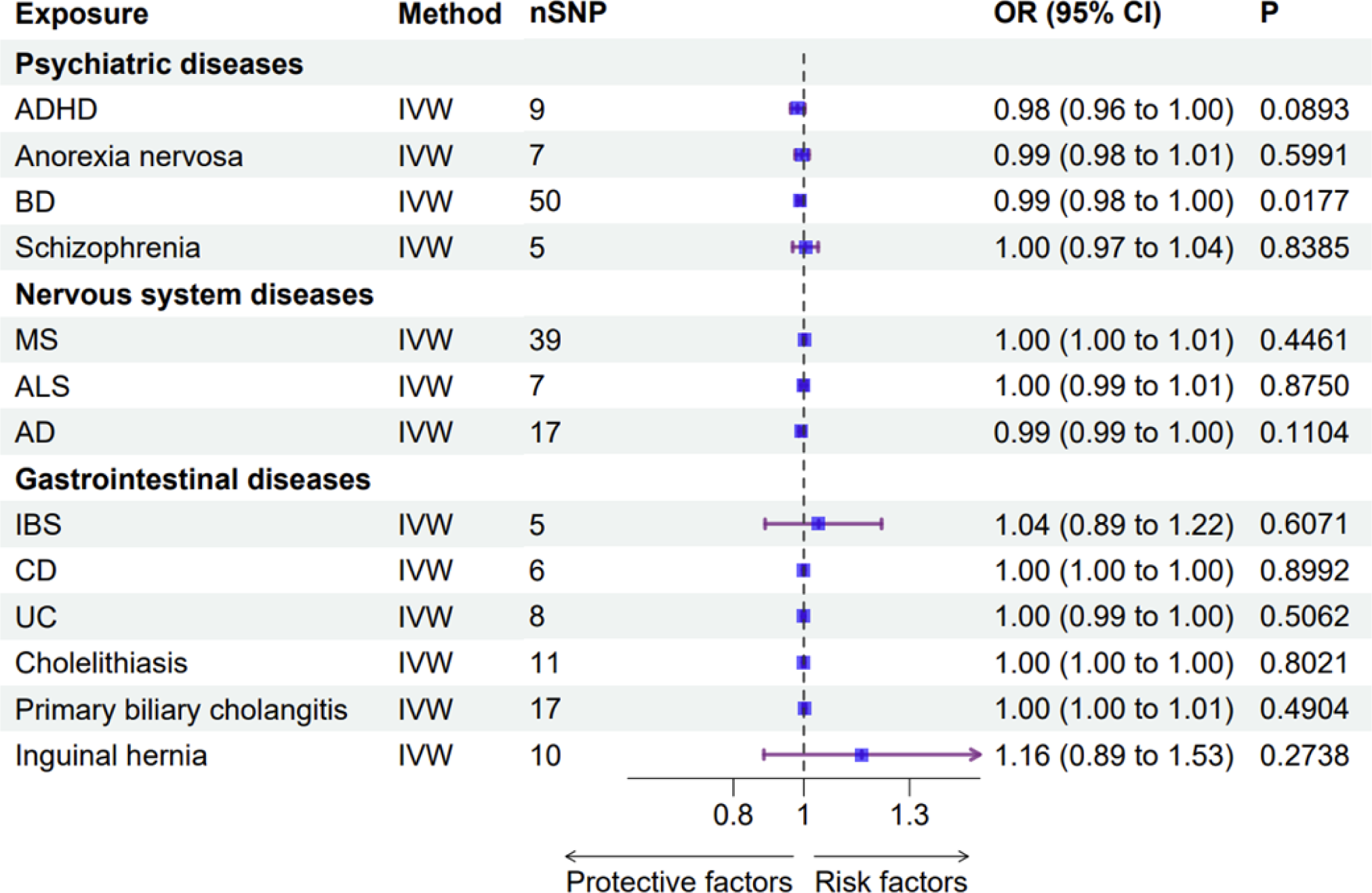
MR estimates for the effect of 42 diseases on “ever smoked”. Estimates were derived by the inverse-variance weighted method. Reported P-values were adjusted for multiple testing using the Benjamini-Hochberg procedure. The solid lines represent 95% CI; CI, confidence interval; OR, odds ratio. ADHD, attention deficit hyperactivity disorder; BD, bipolar disorder; MS, multiple sclerosis; ALS, amyotrophic lateral sclerosis; AD, Alzheimer’s disease; IBS, irritable bowel syndrome; CD, Crohn’s disease; UC, ulcerative colitis.

## Discussion

This investigation presents the most extensive compilation of publicly accessible GWAS data, enabling a comprehensive characterization of the genetic associations, pleiotropy and causality underlying smoking and neuropsychiatric and gastrointestinal diseases. Notably, our findings unveil a significant genetic correlation between smoking and GRED. Moreover, our research identifies six diseases in which smoking exhibits a causal relationship.

In this study, we report, for the first time, that among the 42 neuropsychiatric and gastrointestinal diseases examined, only GERD exhibits a significant genetic association with smoking. This finding aligns with previous epidemiological evidence suggesting a connection between smoking and GRED (23–25). The genetic correlation between smoking and GRED suggests the presence of shared genetic factors, indicating potential associations between the two. However, further investigation is required to determine the precise mechanisms underlying these relationships. Our pleiotropy analysis using MTAG revealed 358 SNPs jointly associated with both smoking and GRED. However, we found no evidence supporting a significant causal relationship between smoking and GRED in our MR analysis after horizontal pleiotropy and heterogeneity tests. This study boasts a substantially larger sample size for GERD, encompassing 129,080 cases and 473,524 controls, totaling 602,604 samples. This heightened sample size enhances the statistical power of our results, rendering this study’s conclusions more reliable and compelling in refuting the previous finding(25) of a causal effect of smoking on GERD (71,522 cases and 261,079 controls) and supporting the notion that the genetic association is more likely attributable to pleiotropy.

Furthermore, we observed a potential genetic link between smoking and psychiatric disorders such as schizophrenia, which corroborates the findings of previous studies(26,27). It is noteworthy that both our study, conducted within a European population, and previous studies conducted in East Asian populations did not find a significant causal relationship between smoking and schizophrenia(28). Moreover, recent research suggests that the potential genetic association between smoking and schizophrenia may stem from a local genetic correlation at 15q25(29). This discovery contributes to the mounting evidence supporting a genetic link between smoking and schizophrenia arising from specific loci rather than causal effects.

Additionally, pathway clustering analysis revealed that the 513 pleiotropic genes shared between smoking and the 42 diseases were enriched for pathways related to growth regulation and synaptic function. This enrichment suggests that smoking may be implicated in disease development by influencing neural and developmental processes. The enriched neurodevelopmental pathways imply that smoking could disrupt crucial growth regulators during neural maturation, thereby affecting disease risk. The synaptic pathways indicate that smoking might influence neurotransmitter signaling and synaptic plasticity, thereby impacting disease pathogenesis. By connecting smoking-associated genes to established disease pathways, our integrative analysis yields new biological insights distinct from previous observational studies. Nonetheless, experimental validations are necessary to confirm the functional roles of the identified genes and pathways in mediating the relationships between smoking and diseases. Our MR analysis provides compelling evidence of causal relationships between smoking and several diseases, including UC, IBD, gastritis, PTSD, cholelithiasis, and epilepsy. These findings underscore the unique strength of our study in comparison to previous research. Consistent with prior studies, our MR analysis verifies the causal relationship between smoking and cholelithiasis, IBD, and gastritis(30). Furthermore, our results align with a meta-analysis reporting an increased risk of gallbladder disease associated with smoking(31). However, our study diverges from previous research by furnishing robust genetic evidence that smoking does not causally affect gastric and duodenal ulcers, irritable bowel syndrome, or CD. Our MR analysis, with its larger sample size and rigorous statistical corrections, offers novel genetic insights into the causal effects of smoking on diseases. In contrast, we did not detect significant causal effects of the diseases on smoking after multiple testing correction. These null associations indicate that while smoking may influence disease risk, the reverse direction of disease liability contributing to smoking initiation is not supported by genetic evidence. Further studies are warranted to replicate these findings and unravel the biological mechanisms underlying the identified causal effects.

Several limitations merit attention in this study. First, the GWAS data primarily pertain to European ancestry populations and may not be fully generalizable to other ethnic groups. Future investigations involving more diverse populations are imperative to validate the findings. Second, the causal inferences derived from MR analysis require further experimental validation to elucidate the specific molecular mechanisms. In addition, the GWAS samples for smoking and diseases may have partial overlap, which could introduce bias and lead to inflated causal associations. Although we endeavored to maximize the independence between exposure and outcome samples during MR analysis, it is crucial to consider the potential sample overlap when interpreting the causal effects. Additionally, the study may be subject to limitations inherent to the statistical methodology, making it challenging to determine weaker causal relationships between smoking and certain diseases. Our findings call for further research into biological pathways to comprehensively delineate the causality between smoking and various diseases, ultimately advancing precision medicine. Despite these limitations, our study offers comprehensive insights into the potential causal effects of smoking on a wide range of disease endpoints within both the neuropsychiatric and gastrointestinal systems. Further rigorous investigations in the future can aid in validating and expanding upon the findings presented here.

## Conclusions

Overall, by integrating multiple genetic approaches, this study unveils the shared genetics and potential pathogenic effects linking smoking to neuropsychiatric and gastrointestinal diseases, providing crucial insights for subsequent research in this domain. Moreover, these findings hold promise for advancing clinical translational applications that aim to elucidate the intricate relationships between smoking and various diseases.

## Data Availability

All relevant data are within the manuscript and its Supporting Information files.

## List of Abbreviations

MR: Mendelian randomisation
GWAS: Genome-wide association study
SNP: Single-nucleotide polymorphisms
GRED: Gastroesophageal reflux disease
PGC: The Psychiatric Genomics Consortium
LDSC: The linkage disequilibrium score regression
MTAG: Multitrait analysis of GWAS
FDR: False discovery rate
GO: Gene Ontology
IVW: Inverse variance weighted
BH: Benjamini‒Hochberg
OR: Odds ratio
CI: Confidence interval
UC: Ulcerative colitis
IBD: Inflammatory bowel disease
PTSD: Post-traumatic stress disorder
CD: Crohn’s disease
ADHD: Attention-deficit/hyperactivity disorder
MDD: Major depressive disorder
BD: Bipolar disorder

## Ethics approval and consent to participate

Not applicable.

## Consent for publication

Not applicable.

## Availability of data and materials

The GWAS datasets supporting the conclusions of this article are available in the GWAS Catalog (https://www.ebi.ac.uk/gwas/), the Psychiatric Genomics Consortium (https://www.med.unc.edu/pgc/) and the MRC-IEU OpenGWAS database (https://gwas.mrcieu.ac.uk/). The Migraine GWAS dataset provided by Hautakangas et al.(32) can be obtained by contacting the International Headache Genetics Consortium. Data analysis was performed using R version 4.2.2. LDSC (https://github.com/bulik/ldsc/), MTAG (https://github.com/JonJala/mtag/) and MAGMA(http://ctglab.nl/software/magma) were used for analysis, and Metascape (https://metascape.org/) was used for gene ontology enrichment analysis. This study was not preregistered.

## Contributions

JCL, GHZ and BL designed and supervised the study, while JYX performed the majority of the analysis and drafted the manuscript. BL provided mentorship, and all authors contributed to the study, interpreted the findings, and reviewed the manuscript. The final version of the manuscript was approved by all authors before publication.

## Acknowledgements

We are grateful for resources from the High-Performance Computing Centre of Central South University and we would like to thank the International Headache Genetics Consortium for providing us with the migraine GWAS summary data, the GWAS Catlog and the PGC and the MRC-IEU OpenGWAS database for providing other GWAS datasets.

## References

1. Cornelius ME, Loretan CG, Jamal A, Lynn BCD, Mayer M, Alcantara IC, et al. Tobacco Product Use Among Adults – United States, 202. 2023;72(18).

2. Kamceva G, Arsova-Sarafinovska Z, Ruskovska T, Zdravkovska M, Kamceva-Panova L, Stikova E. Cigarette Smoking and Oxidative Stress in Patients with Coronary Artery Disease. Open Access Maced J Med Sci. 2016 Oct 28;4(4):636–40.

3. Barkhuizen W, Dudbridge F, Ronald A. Genetic overlap and causal associations between smoking behaviours and mental health. Sci Rep. 2021 Jul 21;11(1):14871.

4. Papoutsopoulou S, Satsangi J, Campbell BJ, Probert CS. Review article: impact of cigarette smoking on intestinal inflammation—direct and indirect mechanisms. Alimentary Pharmacology & Therapeutics. 2020;51(12):1268–85.

5. Winsvold BS, Harder AVE, Ran C, Chalmer MA, Dalmasso MC, Ferkingstad E, et al. Cluster Headache Genomewide Association Study and Meta-Analysis Identifies Eight Loci and Implicates Smoking as Causal Risk Factor. Ann Neurol. 2023 Jul 24;10.1002/ana.26743.

6. Vandebergh M, Goris A. Smoking and multiple sclerosis risk: a Mendelian randomization study. J Neurol. 2020 Oct;267(10):3083–91.

7. Larsson SC, Carter P, Kar S, Vithayathil M, Mason AM, Michaëlsson K, et al. Smoking, alcohol consumption, and cancer: A mendelian randomisation study in UK Biobank and international genetic consortia participants. PLoS Med. 2020 Jul 23;17(7):e1003178.

8. Larsson SC, Mason AM, Bäck M, Klarin D, Damrauer SM, Million Veteran Program, et al. Genetic predisposition to smoking in relation to 14 cardiovascular diseases. European Heart Journal. 2020 Sep 14;41(35):3304–10.

9. Larsson SC, Burgess S. Appraising the causal role of smoking in multiple diseases: A systematic review and meta-analysis of Mendelian randomization studies. eBioMedicine. 2022 Aug;82:104154.

10. Cryan JF, O’Riordan KJ, Cowan CSM, Sandhu KV, Bastiaanssen TFS, Boehme M, et al. The Microbiota-Gut-Brain Axis. Physiological Reviews. 2019 Oct 1;99(4):1877–2013.

11. Gong W, Guo P, Li Y, Liu L, Yan R, Liu S, et al. Role of the Gut-Brain Axis in the Shared Genetic Etiology Between Gastrointestinal Tract Diseases and Psychiatric Disorders: A Genome-Wide Pleiotropic Analysis. JAMA Psychiatry. 2023 Apr 1;80(4):360.

12. Margolis KG, Cryan JF, Mayer EA. The Microbiota-Gut-Brain Axis: From Motility to Mood. Gastroenterology. 2021 Apr;160(5):1486–501.

13. Mappin-Kasirer B, Pan H, Lewington S, Kizza J, Gray R, Clarke R, et al. Tobacco smoking and the risk of Parkinson disease: A 65-year follow-up of 30,000 male British doctors. Neurology. 2020 May 19;94(20):e2132–8.

14. Botteri E, Borroni E, Sloan EK, Bagnardi V, Bosetti C, Peveri G, et al. Smoking and Colorectal Cancer Risk, Overall and by Molecular Subtypes: A Meta-Analysis. Am J Gastroenterol. 2020 Dec;115(12):1940–9.

15. Schizophrenia Working Group of the Psychiatric Genomics Consortium, Bulik-Sullivan BK, Loh PR, Finucane HK, Ripke S, Yang J, et al. LD Score regression distinguishes confounding from polygenicity in genome-wide association studies. Nat Genet. 2015 Mar;47(3):291–5.

16. Turley P, Walters RK, Maghzian O, Okbay A, Lee JJ, Fontana MA, et al. Multi-trait analysis of genome-wide association summary statistics using MTAG. Nat Genet. 2018 Feb;50(2):229–37.

17. Zeng R, Jiang R, Huang W, Wang J, Zhang L, Ma Y, et al. Dissecting shared genetic architecture between obesity and multiple sclerosis. eBioMedicine. 2023 Jul;93:104647.

18. de Leeuw CA, Mooij JM, Heskes T, Posthuma D. MAGMA: generalized gene-set analysis of GWAS data. PLoS Comput Biol. 2015 Apr 17;11(4):e1004219.

19. Zhou Y, Zhou B, Pache L, Chang M, Khodabakhshi AH, Tanaseichuk O, et al. Metascape provides a biologist-oriented resource for the analysis of systems-level datasets. Nat Commun. 2019 Apr 3;10(1):1523.

20. Yu G, Wang LG, Han Y, He QY. clusterProfiler: an R Package for Comparing Biological Themes Among Gene Clusters. OMICS: A Journal of Integrative Biology. 2012 May;16(5):284–7.

21. Burgess S, Butterworth A, Thompson SG. Mendelian Randomization Analysis With Multiple Genetic Variants Using Summarized Data. Genet Epidemiol. 2013 Nov;37(7):658–65.

22. Lee CH, Cook S, Lee JS, Han B. Comparison of Two Meta-Analysis Methods: Inverse-Variance-Weighted Average and Weighted Sum of Z-Scores. Genomics Inform. 2016;14(4):173.

23. Sharma P. Barrett Esophagus: A Review. JAMA. 2022 Aug 16;328(7):663.

24. Maret-Ouda J, Markar SR, Lagergren J. Gastroesophageal Reflux Disease: A Review. JAMA. 2020 Dec 22;324(24):2536.

25. Yuan S, Larsson SC. Adiposity, diabetes, lifestyle factors and risk of gastroesophageal reflux disease: a Mendelian randomization study. Eur J Epidemiol. 2022 Jul;37(7):747–54.

26. Hahad O, Daiber A, Michal M, Kuntic M, Lieb K, Beutel M, et al. Smoking and Neuropsychiatric Disease—Associations and Underlying Mechanisms. IJMS. 2021 Jul 6;22(14):7272.

27. Hartz SM, Horton AC, Hancock DB, Baker TB, Caporaso NE, Chen LS, et al. Genetic correlation between smoking behaviors and schizophrenia. Schizophrenia Research. 2018 Apr;194:86–90.

28. Chen J, Chen R, Xiang S, Li N, Gao C, Wu C, et al. Cigarette smoking and schizophrenia: Mendelian randomisation study. Br J Psychiatry. 2021 Feb;218(2):98–103.

29. Gerring ZF, Thorp JG, Gamazon ER, Derks EM. A Local Genetic Correlation Analysis Provides Biological Insights Into the Shared Genetic Architecture of Psychiatric and Substance Use Phenotypes. Biological Psychiatry. 2022 Oct;92(7):583–91.

30. Yuan S, Chen J, Ruan X, Sun Y, Zhang K, Wang X, et al. Smoking, alcohol consumption, and 24 gastrointestinal diseases: Mendelian randomization analysis. eLife. 12:e84051.

31. Aune D, Vatten LJ, Boffetta P. Tobacco smoking and the risk of gallbladder disease. Eur J Epidemiol. 2016 Jul;31(7):643–53.

32. Hautakangas H, Winsvold BS, Ruotsalainen SE, Bjornsdottir G, Harder AVE, Kogelman LJA, et al. Genome-wide analysis of 102,084 migraine cases identifies 123 risk loci and subtype-specific risk alleles. Nat Genet. 2022 Feb;54(2):152–60.

